# Practitioner, patient and public views on the acceptability of Mobile Stroke Units in England and Wales: a mixed methods study

**DOI:** 10.1101/2024.08.26.24312612

**Authors:** L. Moseley, P. McMeekin, C. Price, L. Shaw, M. Allen, G.A. Ford, M. James, A. Laws, S. McCarthy, G McClelland, L.J. Park, K. Pearn, D. Phillips, P. White, D. Wilson, J. Scott

## Abstract

**Background:** Evidence for Mobile Stroke Units (MSUs) demonstrates that onset to treatment times for intravenous thrombolysis can be reduced and access to mechanical thrombectomy might be improved. Despite growing use of MSUs internationally, to date there have been no studies in NHS England and NHS Wales exploring the acceptability of MSUs to clinicians, patient and public representatives and other key stakeholders, which are important when considering potential feasibility and implementation.

**Methods:** This study used a mixed methods design with a cross-sectional survey and qualitative workshops and interviews between October 2023 to May 2024. Survey data were collected from clinicians involved in emergency stroke care. Qualitative data involved clinical and non-clinical professionals involved in stroke care alongside patient and public representatives with experience of stroke. Survey data were descriptively analysed while content analysis was used on open-ended questions. Qualitative data were thematically analysed, prior to triangulation using a convergent coding matrix.

**Results:** The study results, drawn from 25 respondents to the survey and 21 participants in qualitative workshops, found that almost all participants had positive affective attitudes to the concept of MSUs. However, several key areas of concern were identified that need to be addressed prior to implementing MSUs. These concerns included how MSUs would be staffed; whether and how telemedicine could contribute; the types of economic impacts; extent to which triage systems could accurately identify stroke patients for MSUs to attend; where the base location and geographic coverage of MSUs should be, the impact of MSUs on equitable access to stroke care, and how to improve public awareness of MSUs.

**Conclusion:** Whilst MSUs are mostly acceptable to key stakeholders, numerous areas of concern need to be addressed prior to MSU implementation. We recommend further research to address these issues prior to implementation in the NHS.

## Introduction

Stroke is the leading cause of disability and second highest cause of mortality worldwide[1]. There are two main types: 85% are ischaemic as a result of blood clot causing a blockage in the brain, and 15% are haemorrhagic, resulting from bleeding within the brain. In both cases there is usually a rapid onset of neurological symptoms, which can be minimised through early medical assessment and treatment. Ischaemic stroke can be treated in up to 20% cases with intravenous thrombolysis (IVT) which restores blood flow to brain tissue and produced a measurable reduction in stroke-related disability for approximately one third of patients[2]. For patients with ischaemic stroke due to large vessel occlusion (LVO), a large blood clot is located in a main vessel supplying the brain and the prognosis is particularly poor despite IVT. However, for selected patients the clot can be surgically removed by an interventional neuroradiologist at centres with appropriate specialist facilities. This mechanical thrombectomy (MT) procedure re-opens the artery and significantly improves outcomes for one in three patients[3], but to access treatment many have to be transferred from their local hospital to a regional centre. Currently the main treatments for the less common scenario of haemorrhagic stroke includes controlling high blood pressure to slow the bleeding, correction of blood clotting abnormalities and surgical intervention for highly selected patients[4]. In order to decide which patients with suspected acute stroke in the community are suitable for any of these treatments and to gain maximum benefit, they require emergency assessment by ambulance practitioners and rapid transportation to hospital for brain imaging and specialist review.

Mobile Stroke Units (MSUs) have been evaluated as an approach to bring diagnosis and time-critical stroke treatment to patients instead of transporting them by ambulance to the nearest hospital that can provide IVT. These customised vehicles are equipped with computed tomography (CT) scanners, point of care laboratory equipment, and access to clinical stroke expertise in person or via telemedicine[5]. Randomised controlled trial evidence from outside the United Kingdom (UK), including in the USA, Australia and Europe, show that MSU improve the rate and timeliness of intravenous thrombolysis in urban settings[6–10]. In addition it has been proposed that MSUs with the added technology to perform blood flow imaging (CT angiography) could improve outcomes by directing patients with large vessel occlusion to the nearest facility offering mechanical thrombectomy if this is unavailable locally[11–13], although definitive evidence is still lacking[6–10, 14]. There has also been a report from in the UK that MSU assessment can reduce hospital admissions of ‘stroke mimic’ patients who have more benign conditions that can produce stroke-like symptoms, such as migraine[15]. As clinical trials have consistently reported MSU benefits for IVT delivery, the European Stroke Organisation[16] recommends their implementation but the evidence reflects the settings where trials were hosted and will be context-dependent, varying in relation to infrastructure, demographics, geography, and the costs of standard care[17, 18]. In recent years ambulance response times have decreased[19] and emergency department waiting times have increased[20], likely leading to reductions in access to emergency health care, which would encompass stroke care[21]. As a result, MSUs may be seen to be an attractive policy option for the NHS to consider, though evidence on the barriers and facilitators to their implementation is lacking in the UK, as well as how evidence from other countries would translate to a UK setting.

Despite evidence and recommendations favouring the expansion of MSU, there is limited research exploring professional and public perspectives. As part of a process evaluation for a trial of MSUs in Australia, Bagot et al.[22, 23] identified many practical implementation challenges but did not explore wider acceptability, such as attitudes of clinicians and the public. Without acceptability amongst key stakeholders, any potentially beneficial intervention may not be feasible or deployed as intended[24] [25], and may require additional workarounds which impact upon the effectiveness and cost-effectiveness. Acceptability has had an inconsistent definition throughout the literature, with varying approaches and measures. The Theoretical Framework of Acceptability (TFA)[26] describes multiple components that form and explain overall acceptability of innovations and interventions in healthcare. Informed by the TFA, this paper aims to examine affective attitudes of relevant stakeholders to MSUs, including economic and equity considerations, to inform future considerations for their implementation in England and Wales. Consideration of acceptability is essential and as such the consideration of acceptability of a proposed new intervention is the first stage of the framework for development of evaluation of complex interventions guidance by the Medical Research Council (MRC)[24].

## Methods

Ethical approval was provided via Northumbria University ethics online system (reference: 4117). The study was deemed by the Health Research Authority (HRA) to not require HRA approval. All participants gave written consent prior to any data collection which was returned by email and stored securely by the researcher.

### Study Design

A concurrent mixed methods study design consisting of a quantitative cross-sectional survey and qualitative workshop, focus group and semi-structured interviews. Data were collected in the context of a wider study that is aiming to co-design economic models which consider the cost implications and potential cost benefits of MSUs, and quantitative models that consider impacts of MSUs on treatment times[27].

### Participants

Survey participants were recruited from attendees at a nationally advertised stroke clinical education and service improvement meeting held in York, England on 20^th^ October 2023. The event was attended by a range of health care professionals with direct clinical experience of emergency stroke care.

Participants for qualitative data collection were recruited using a combination of sampling techniques including purposive, opportunistic and snowball sampling between 11^th^ September 2023 and 2^nd^ July 2024. This consisted of identifying relevant stakeholder groups (stroke clinicians (physicians, nurses), ambulance service staff, stroke patients or carers of people who have experienced a stroke), with participants identified via existing networks of the research team.

Patient and public participants were recruited via the Stroke Association, a charitable organisation that supports people to rebuild their lives after stroke.

### Data collection

Survey questions were designed by a team of researchers, clinicians and public representatives (co-authors) to obtain information about participant characteristics; prior knowledge of MSUs; dispatch and operational factors; geographical and equity factors; funding and whether they viewed MSUs as feasible in the current NHS. The survey was accessed online using a QR code or url link with responses captured via a mixture of 5-point Likert scales, single-response multiple choice, and open-ended questions. Qualitative data were collected using a topic guide which included discussions about the prior knowledge of MSUs; whether they were for or against the implementation of MSUs and why; concerns around feasibility and implementation challenges. All qualitative data were recorded using an encrypted recorder and transcribed verbatim.

### Analysis

Survey data were descriptively analysed using Microsoft Excel 365 (Microsoft Corporation) and are presented in narrative and tabular formats. Open-ended survey questions were analysed using content analysis and presented based on overall categories. Qualitative data were thematically analysed[28] focusing specifically on people’s views of MSUs. Nvivo Version 12 (Lumivero) was used to aid analysis. One team member (LM) initially coded the data into themes with another team member (JS) independently analysing a sample of 20% of the transcripts. LM and JS then refined the themes together before discussion with the wider research team. Mixed methods data were then triangulated by LM and JS using a convergent coding matrix,[29] examining the (dis)agreements and silences across the survey and qualitative data. In the sections that follow and building on this commitment to data integration, we have reported qualitative and quantitative data concurrently.

## Results

Of 110 people invited to complete the survey, twenty-five (23%) did so. Twenty-one people contributed qualitative data in a workshop (n=10), semi-structured interviews (n=6) and a focus group (n=5). The qualitative participants were recruited separately from survey participants except one participant who participated in both the survey and the qualitative aspect. All participant characteristics, for the survey and qualitative data, are shown in Table 1. Survey findings are reported in Table 2. For the interview results, eight themes were developed that captured stakeholders’ affective attitudes to MSUs, including anticipated implementation challenges, which are shown in figure 1.

**Table 1:** Participant characteristics.

| Survey |  |
| --- | --- |
| Characteristic | Number of respondents |
| **Gender** |  |
| Female, n (%) | 10 (40) |
| Male, n (%) | 15 (60) |
| **Professional background** | **Number of respondents** |
| Consultant stroke physician, n (%) | 9 (36) |
| Junior doctor, n (%) | 8 (32) |
| Nurse stroke practitioner, n (%) | 4 (16) |
| Other: Speciality registrar, n (%) | 2 (8) |
| Other: Physician associate, n (%) | 1 (4) |
| Other: Advanced clinical practitioner, n (%) | 1 (4) |
| **Mean length of time qualified (range; standard deviation)** | 8.5 years (1 to 23; 6.2)* |
| **Work exclusively in stroke care** | **Number of respondents** |
| Yes, n (%) | 16 (64) |
| No, n (%) | 9 (36) |
| **Employment Location** | **Number of respondents** |
| North East and North Cumbria, n (%) | 12 (48) |
| East Midlands, n (%) | 3 (12) |
| East of England (North), n (%) | 2 (8) |
| Cheshire and Mersey, n (%) | 1 (4) |
| East of England (South), n (%) | 1 (4) |
| *Greater Manchester, n (%)* | 1 (4) |
| *Kent and Medway, n (%)* | 1 (4) |
| *South Yorkshire, n (%)* | 1 (4) |
| *Swindon & Wiltshire, Bristol, North Somerset & South Gloucestershire and Somerset, n (%)* | 1 (4) |
| *West Midlands, n (%)* | 1 (4) |
| *West Yorkshire and Harrogate, n (%)* | 1 (4) |
| **Previously heard of Mobile Stroke Units** | **Number of respondents** |
| *Yes, n (%)* | 24 (96) |
| *No, n (%)* | 1 (4) |
| **Workshops, focus groups and interviews** |  |
| **Characteristic** | **Number of Participants** |
| **Gender** |  |
| *Female, n (%)* | 9 (43) |
| *Male, n (%)* | 12 (57) |
| **Primary background** |  |
| *Patient and public involvement representative, n (%)* | 11 (52) |
| *Stroke consultant, n (%)* | 5 (24) |
| *Ambulance service staff\*\*\*, n (%)* | 5 (24) |

\* n=24 (96% of participants); one participant did not provide their length of time qualified.
\*\* including paramedics, service lead and medical director.

**Table 2:** Summary of survey responses.

| **Question** | **Number of Respondents (%)** |
| --- | --- |
| ***Which service should operate MSUs on a daily basis?*** |  |
| *Hospital service, n (%)* | 9 (36) |
| *Ambulance service, n (%)* | 9 (36) |
| *Unsure, n (%)* | 4 (16) |
| *Other (all respondents who selected other specified a joint approach), n (%)* | 3 (12) |
| ***Where should the MSU be based physically between calls?*** | **Number of Respondents (%)** |
| *Ambulance station, n (%)* | 8 (32) |
| *Regional stroke hospital, n (%)* | 7 (28) |
| *Local stroke unit, n (%)* | 6 (24) |
| *Ambulance headquarters, n (%)* | 2 (8) |
| *Unsure, n (%)* | 2 (8) |
| ***Which geographical area(s) should the MSU primarily operate in?*** | **Number of Respondents (%)** |
| *All areas, n (%)* | 14 (56) |
| *Rural, n (%)* | 10 (40) |
| *Suburban, n (%)* | 1 (4) |
| *Central urban, n (%)* | 0 (0) |
| *Unsure, n (%)* | 0 (0) |
| ***Should a MSU, where practically possible, respond to all suspected stroke emergency (999) calls?*** | **Number of Respondents (%)** |
| *No, n (%)* | 12 (48) |
| *Yes, n (%)* | 11 (44) |
| *Unsure, n (%)* | 2 (8) |
| ***Which scenarios should a MSU not respond to emergency (999) calls?*** | **Number of respondents (% of all answers)\*** |
| *Low GCS/clinically unstable/deteriorating rapidly, n (%)* | 3 (27) |
| *Patients who can be transported to a stroke unit quicker than MSU can respond, n (%)* | 2 (18) |
| *High probability of a stroke mimic, n (%)* | 2 (18) |
| *Before paramedics have responded and requested MSU, n (%)* | 2 (18) |
| *Ambiguous symptomology, n (%)* | 1 (9) |
| *Unsure, n (%)* | 1 (9) |
| **What level of risk of an adverse event would a patient be at when receiving stroke treatment on a MSU compared to in-hospital treatment at a specialist stroke centre?** | **Number of Respondents (%)** |
| *Same risk, n (%)* | 13 (52) |
| *Greater risk, n (%)* | 8 (32) |
| *Lesser risk, n (%)* | 2 (8) |
| *Unsure, n (%)* | 2 (8) |
| **Would you support a MSU if one was available in your own services?** | **Number of Respondents (%)** |
| *Yes, occasionally as part of my current role, n (%)* | 14 (56) |
| *Yes, exclusively, n (%)* | 8 (32) |
| *No, n (%)* | 3 (12) |
| **To what extent:** | **Median (IQR)<sup>†</sup>** |
| *Would MSUs increase or decrease equitable access to stroke care?* | 4 (1) |
| *Do you feel you understand what a MSU does?* | 4 (1) |
| *Do you think MSUs would disrupt existing emergency stroke care pathways?* | 2 (2) |
| *Do you feel confident that stroke care can be delivered on a MSU via telemedicine instead of having a stroke physician physically present?* | 3 (2) |
| *Would MSUs be beneficial in the NHS in England and Wales?* | 4 (2) |
| *Should MSUs be prioritised for funding in consideration of other financial challenges in the NHS?* | 3 (1) |
| *Should MSUs be prioritised for funding in consideration of other developments in stroke care?* | 3 (1) |
| **Overall:** | **Median (IQR)<sup>†</sup>** |
| *MSUs appear feasible to me in the current NHS* | 2 (2) |
| *MSUs are acceptable to me in the current NHS* | 3 (2) |
| **Content analysis of challenges and other considerations** | **Number of respondents (% of all answers\*)** |
| *Workforce: staffing of MSU (staff mix and availability of staff, n (%))* | 13 (27) |
| *Cost of the MSU, n (%)* | 12 (25) |
| *Evidence base: need for pilot work, n (%)* | 5 (10) |
| *Workforce: training and skill, n (%)* | 5 (10) |
| *Equity of access and geography, n (%)* | 4 (8) |
| *Technical: reliable access to imaging, n (%)* | 3 (6) |
| *Embedded in stroke pathway, n (%)* | 2 (4) |
| *Technical: telehealth, n (%)* | 2 (4) |
| *Workforce: collaborative working, n (%)* | 2 (4) |

GCS=Glasgow Coma Score. IQR=Interquartile range. MSU=mobile stroke unit. NHS = National Health Service.
\* optional free-text question, respondents could provide no response, single response or multiple responses.
† five-point Likert scale, 1=not at all, 5=completely.

**Fig 1:**
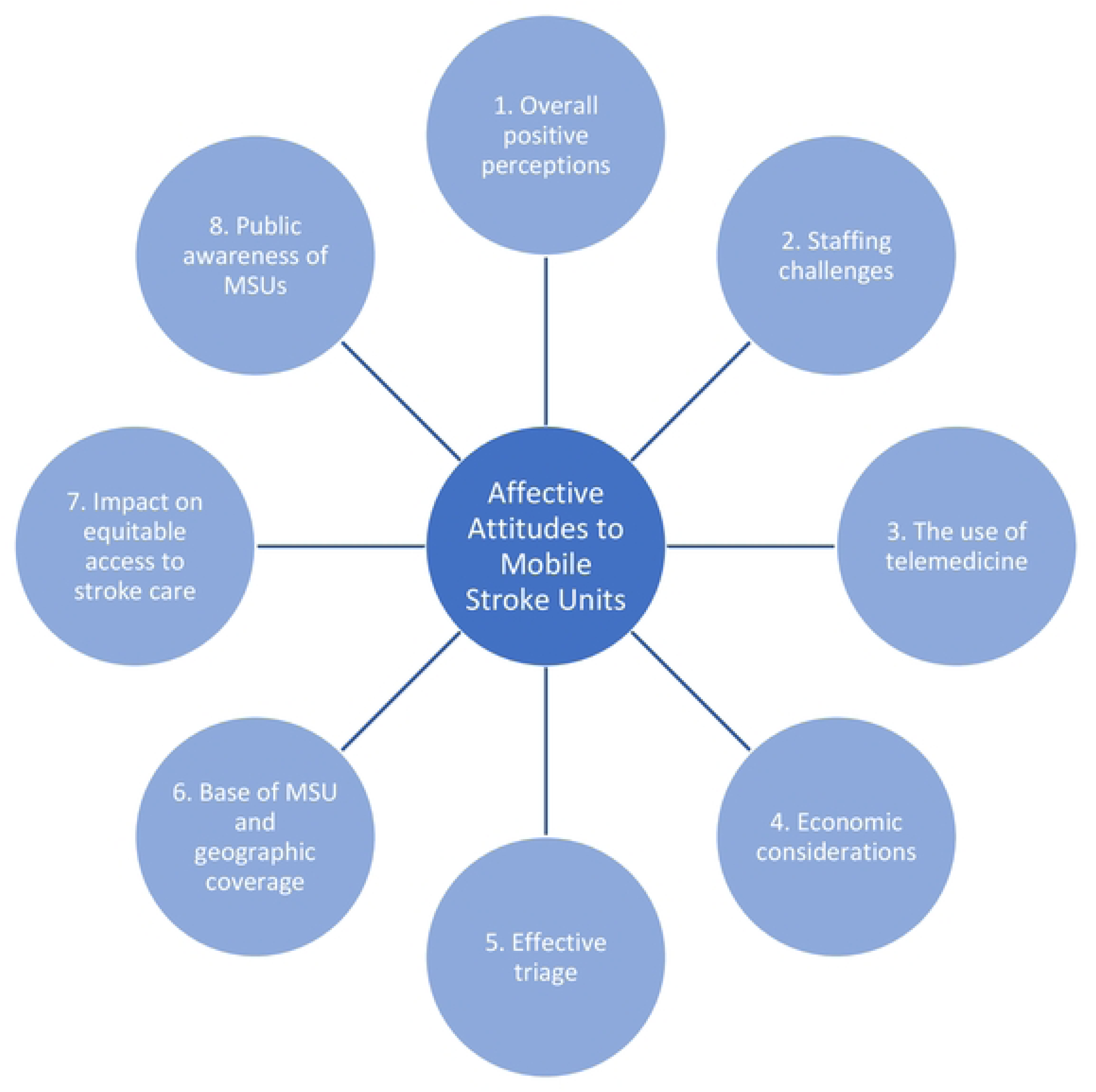
Summary of themes showing stakeholders’ affective attitudes to Mobile Stroke Units.

### Theme 1: Overall perception of MSUs

Survey data showed a generally supportive view on MSUs with the majority of participants stating they would work on an MSU either exclusively or as part of their existing role (n=22, 88%), but with reservations about their acceptability in National Health Service (NHS) England and NHS Wales, with a median score of 3 on 1-5 Likert scale (with 1 being not at all acceptable and 5 being completely acceptable). Qualitative data also highlighted a positive overall view of the potential of MSUs, when participants were initially asked about their thoughts on MSUs as illustrated by this quote from an ambulance service member of staff:

> *“So, I love it. I think it’s a brilliant concept. It’s bringing the CT scanner to the patient in the community, which ultimately is going to reduce the [thrombolysis] times and we know that the earlier patients get treatment then the better their prognosis, the less rehabilitation they need and the less disability they’re left with.” (Participant 16, ambulance service staff, semi-structured interview)*

Only two (9.5%) participants expressed a less favourable initial opinion, which was based upon anticipated implementation challenges including those related to geographical equity. This is summed up by a stroke consultant:

> *“I’ve heard a lot of the tales of mobile stroke units and I will admit to being a cynic. I’m interested to have some chats about how we use them in more challenging areas of the country and populations that maybe a bit more rural…” (Participant 7, stroke consultant, workshop)*.

Concerns about feasibility may have impacted survey participants views on acceptability, which may explain the difference between high acceptability in the qualitative data compared with a mixed response in the survey data. Survey data showed participants did not feel they were currently feasible in NHS England and NHS Wales (median=2, on a 1-5 Likert scale with 1 being not at all feasible and 5 being completely feasible) which is likely to influence views on acceptability.

### Theme 2: Staffing challenges

Staffing, including associated workforce considerations, was the most reported concern highlighted by survey participants. This was also a notable concern for qualitative participants with the current shortage of stroke physicians within the NHS being highlighted, and that MSUs would inevitably place further strain on this already limited resource, including reducing the number of available stroke clinicians available in hospitals. This was summed up by a member of ambulance service staff:

> *“the model of having to have a doctor onboard and a neurologist onboard […] has obviously got its challenges because there are not many…at the moment.” (Participant 16, ambulance service staff, semi-structured interview)*

The combination of staff required on MSU was mentioned as frequently as staffing availability within the free text area of the survey and was also a key consideration in the qualitative data. There were mixed views on what the optimum staffing mix would be, but all agreed that regardless of professional background, MSU staff would need to be experienced in providing stroke care and likely have additional training. This was summarised by a member of ambulance service staff.

> *“Your staffing configuration involves senior clinicians from both or any services that work on it, we wouldn’t ever put a newly qualified paramedic on or a relatively newly qualified doctor.” (Participant 3, ambulance service staff, workshop)*

### Theme 3: The use of telemedicine

The use of telemedicine, where patient scans could be shared with a physician remotely to facilitate diagnosis and treatment decisions, was discussed as a potential opportunity to overcome concerns about stroke physician shortages. There was overall favourable support for telemedicine amongst qualitative participants, however survey data with clinicians showed that those participants were unsure about the use (median=3, neither confident nor unconfident). Qualitative participants, particularly clinicians, raised some concerns about telemedicine usage, mainly the need for mobile data signal which may not be available in some rural areas of the country. This would hinder the ability to contact a stroke clinician and transmit scan images. Despite these concerns it was felt telemedicine could be viable and that the impact of Covid-19, which necessitated the use of higher levels of telecommunication within healthcare, had given a more favourable view of the use of telemedicine, including amongst patients. For example, a PPI representative contextualised how patients and the public are now used to telemedicine:

> *“… post-Covid, I think [telemedicine] is a different beast. And I think we are all much more comfortable with remote advice, not doing face-to-face” (Participant 19, PPI representative, focus group)*

### Theme 4: Economic considerations

The cost of MSUs was the second highest concern reported in free text survey comments. Qualitative participants, including those with experience of working on ambulance services, also raised that the running costs of MSUs could be significant and of concern. Despite not being presented with any economic analysis, they felt it was likely that this would offset costs elsewhere, including possible direct cost savings from stroke patients who are seen quicker, thus making a quicker recovery, and reducing longer term costs of care required both to health and social care as well as returning to work. Participants also highlighted potential indirect cost savings including reducing the number of people conveyed to emergency departments (EDs), by diverting stroke patients, which would also reduce ambulance waiting times at EDs. Further, MSUs were thought to have the potential to diagnose and treat conditions at home, including stroke mimics and transient ischemic attacks, with appropriate onward referrals, again reducing ED and ambulance service pressures. This was explained by a member of ambulance service staff:

> *“…the MSU is an absolute priority in identifying stroke, but also it’s valuable in identifying those that are not stroke and then reducing the impact on the wider NHS further…those patients that are identified as non-stroke may not be conveyed.” (Participant 5, ambulance service staff, workshop)*

The second consideration regarding the costs of MSUs is whether that money would be better invested elsewhere. Survey data showed that participants were unsure whether MSUs should be prioritised for funding in stroke care (median=3, neither prioritised or not prioritised), and this same uncertainty was shared by qualitative participants. Improvements to rehabilitation services following a stroke and increasing thrombectomy services were highlighted by a PPI representative as a possibility:

> *“…why the money wasn’t put into thrombectomy rather [than] a mobile stroke unit because it’s not available everywhere and it’s nowhere near the target figures…” (Participant 11, PPI representative, workshop)*

### Theme 5: Effective triage

All qualitative participants raised concerns that current triage systems used to dispatch ambulances would unlikely be robust enough to be solely relied upon to dispatch MSUs. This was felt to be an important factor in the acceptability of MSUs including balancing efficiency whilst ensuring MSUs are not underutilised. This was explained by a member of ambulance service staff:

> *“The dispatch system, when you look at it in terms of effectiveness…to identify stokes isn’t brilliant…” (Participant 3, ambulance service staff, workshop)*

Survey data showed some uncertainty around who the MSU would respond to, with 48% (n=12) stating the MSU should not respond to all stroke calls and 44% (n=11) saying that it should. The main reasons given for not responding were conveyance times, patient’s condition, and whether symptoms are suggestive of a mimic. This aligns with qualitative data, which highlighted that current triage systems are unlikely to reliably achieve dispatch to the most appropriate people.

### Theme 6: Base of the MSU and Geographic Coverage

The base location and geographic coverage of the MSU was another area of key uncertainty. Survey respondents gave mixed opinions about whether the MSU should be based at an ambulance station (n=6, 32%), regional stroke hospital (n=8, 28%) or local stroke unit (n=7, 24%). Views were similarly mixed within qualitative data, with the practicality of operating an MSU being a key consideration:

> *“…an ambulance service not only provides care to patients, but it also specialises in fleet management, maintenance, and also specialises in dispatch, which, unless you’re going to do those things, how are you going to keep your MSU up and out on the road?…But then equally…[the ambulance service] don’t directly employ a neurologist…So we would need to be in collaboration with an employer* that had that available resource to work with us.” (Participant 3, ambulance service staff, workshop)

Equally as uncertain was where the MSU would likely operate on a day-to-day basis. Survey data were conflicting, with 56% (n=14) reporting the MSU should cover urban and rural areas, whilst 40% (n=10) reported MSU should focus on rural areas. Exploration of geographical coverage in the qualitative data gave an understanding of the uncertainty. Participants felt that basing the MSU in more rural areas would improve stroke care, and likely outcomes, for those who currently would have long travel times for treatment. However, it was recognised that the MSU would then potentially see a relatively much smaller number of patients, which would likely impact on cost effectiveness. In comparison, MSUs based in urban areas may see a larger number of patients, but this population likely already has relatively better access to stroke care.

### Theme 7: Impact on equitable access to stroke care

The uncertainty in geographical coverage also raised discussion about equitable access to stroke care and the impact of MSUs. Survey data showed participants felt MSUs would improve equitable access (median=4, minor increase). However, qualitative data showed that while MSUs could potentially improve equitable access, because of the previously discussed concerns about placing the MSU in a rural area, where it would have the biggest impact on equitable access, MSUs were considered unlikely to improve equitable access:

> *“One of the factors…is the more people you aim to treat, the more funding you’ll get in future to grow the service…But from a patient perspective, obviously those in more rural areas are more challenged to get to the right place…and the intervention by the [mobile stroke] unit could be more meaningful than…the [MSU] arriving and reducing an urban travel time.” (Participant 2, PPI representative, semi-structured interview)*

### Theme 8: Public awareness of MSUs

Whilst silent within survey data – likely due to the survey being limited to stroke professionals - public awareness of MSUs was raised in the qualitative data. There were concerns around the name Mobile Stroke Unit; participants felt this did not accurately convey the purpose or capabilities of MSUs, instead likening it to vehicles used in public places for health screening. A PPI representative, advised:

> *“To me, a mobile stroke unit is something that sits outside a public library or a festival and people go in and get their blood pressure checked and they talk to someone about their probability of having a stroke and what they can do about it…the name should be stroke ambulance because a unit doesn’t involve any sense of emergency.” (Participant 19, PPI representative, focus group)*

Participants felt that raising awareness of MSUs, including the potential benefits, was a key element of MSUs being acceptable to the wider public. This was summed up by a PPI representative:

> *“…once these units are deployed, people will want to tell their stories…So that builds both a critical awareness and then acceptance that this is really the way forward.” (Participant 2, PPI representative, semi-structured interview)*

## Discussion

This is the first study to examine acceptability of MSUs in NHS England and NHS Wales, drawing on a wide range of relevant stakeholders using mixed methods. Despite participants overall being largely supportive of MSUs, multiple challenges were identified that require addressing prior to possible wider-scale implementation including how to staff the MSU; clarity over what an MSU is and isn’t; whether, and how, telemedicine could contribute; of if/how triage systems could accurately identify stroke for MSUs to attend; the types of impact; where the base location and geographical coverage of MSUs should be and how to improve public awareness of MSUs.

These challenges are similar to those identified in a process evaluation of a trial MSU in Australia[22, 23], suggesting that they are likely to span across different healthcare systems. Broader studies on the organisation of emergency medical systems could provide insight into overcoming the identified challenges, such as the existing use of telemedicine in pre-hospital stroke care[30] and how ambulance locations can be optimised to provide the greatest coverage[31]. There is also a clear perceived trade-off between geographical area covered and the number of patients likely attended, which would likely impact on both clinical and cost effectiveness of MSUs. Previous trials of MSUs have primarily taken place in larger urban areas and responded to patients already in close proximity to large hospitals with the capabilities to treat stroke[6–10]. However, this was a clear source of tension with participants’ equity and economic considerations. Other challenges, specifically improving existing triage systems and addressing structural staffing issues, are likely to be more elusive. For instance, despite a warning to the NHS in 2019 that stroke consultants needed to increase by a third to provide an adequate service[32], staffing continues to be challenging in providing emergency stoke care across existing services, with the Stroke Association recently highlighting that workforce pressures have continued to grow[33]. Triage systems also lack sensitivity and specificity in recognising acute stroke, requiring further development of algorithms[34, 35] or alternative approaches to identifying stroke such as increased point of care testing[36]or video triage[37].

The development of suitable MSU pathways and planned modelling work, including modelling potential cost burdens and benefits and time to treatment models[27], is likely to provide further insight into and options for addressing some of these challenges. Through engagement with stakeholders the pathway development will consider how to overcome challenges in relation to staffing, telemedicine and triage within the NHS. Further exploration of the economic impact, equitable access to stroke care and the geographical location of MSUs will be achieved through co-produced economic and quantitative modelling. In relation specifically to cost-effectiveness, only four studies included in the ESO guidance reported on whether MSUs are cost-effective[17, 18, 38, 39], two of which[18, 38] found MSUs would be cost-saving compared to usual care, and two[17, 39] found cost-effectiveness contingent on notional willingness to pay for a quality-adjusted life year (QALY) or a disability-adjusted life year (DALY), illustrating a lack of evidence and context dependency of cost-effectiveness of economic evaluations[40]. Concerns were also raised in our study about the expenditure required for MSUs when other areas of established stroke care, such as mechanical thrombectomy, require additional resources to increase accessibility to the whole population[41]. Those commissioning MSUs would likely need to factor in how best to utilise limited resources for greatest patient benefit or reduction of inequalities. Future modelling work is unlikely to be able to address all of the concerns and would likely only be explored in future pilots and trials, given the technological and socio-cultural processes involved in the implementation of research-based evidence into practice[42].

### Limitations

It is important to be aware of responder bias for the survey data, as this was a voluntary and anonymous survey at a single national meeting. The survey also has a high geographical representation from the North East of England which may impact on the results. Further, sampling of some qualitative participants may have introduced bias due to already having an interest in MSUs, though scepticism amongst some participants and the identification of multiple challenges suggests that this was minimal.

### Conclusion

Evidence is building around the effectiveness of MSUs for providing more timely treatment for acute ischaemic stroke yet is mainly based on MSUs implemented in single urban areas without consideration for wider-scale implementation and adoption across a whole healthcare system. By examining stakeholders’ views around the acceptability of MSUs, which were largely favourable, we have identified several challenges to implementing MSUs in NHS England and NHS Wales.

## Data Availability

Data cannot be shared publicly because participants did not give consent for data sharing. Data are available from Northumbria University, contact via corresponding author, for researchers who meet the criteria for access to confidential data.

## Acknowledgements

We would like to thank the Stroke Association for helping us to identify participants, and all participants for giving their time.

